# Performance of new national health insurance fund packages (Wekeza, Najali, Timiza) in Kinondoni municipal, Dar es salaam, Tanzania

**DOI:** 10.1101/2024.03.19.24304524

**Authors:** Evangelina C. Chihoma, Mughwira Mwangu

## Abstract

**Background:** Tanzania is one of the countries which has joined the UN efforts of attaining Universal health Coverage (UHC) by 2030 by ensuring that all citizens are able to access health services thereby reducing out of pocket expenditure. As a strategy to achieve UHC, the National Health Insurance Fund (NHIF) introduced new premium packages in September 2019 named TIMIZA, NAJALI and WEKEZA premium packages to accommodate private individuals and their families who are not in the formal employment sector to increase the coverage of citizens who are health insured in the country and to increase their accessibility to quality health services. Since the introduction of these packages no studies have been done to assess their performance. Therefore, this study evaluated the performance of the packages through process evaluation by exploring its level of utilization from September 2019, the acceptability of the packages by the health care providers and client satisfaction towards the services offered.

**Methods:** A descriptive cross-sectional study was conducted in selected NHIF accredited health facilities in Kinondoni Municipal using mixed method of data collection. Secondary data was quantitatively extracted from NHIF enrollment registries using a data abstraction tool to assess the percentage of citizens enrolled from Kinondoni NHIF office from September 2019 to March 2021.In-depth interview was used to evaluate the acceptability of the packages among the health care providers and Clients (beneficiaries) satisfaction towards the quality of services offered. Qualitative data was analyzed by content analysis approach and Microsoft excel for the quantitative data.

**Results:** NHIF has been able to enroll 17,248 members which is only 1% of the total population of people in Kinondoni MC against their set target which was to enroll at least 50% of the total population. Beneficiaries of the packages are unsatisfied and health providers are also unhappy about the packages due to poor orientation, cumbersome referral system and uncertainties in availability of medications and investigations.

**Conclusion and Recommendation:** This evaluation has revealed that NHIF new packages have not been performing well since their introduction in September 2019. They are far from reaching the set target due to challenges which have been brought forward by beneficiaries and health care providers. NHIF need to improve their orientation by educating their clients about every service covered in a scheme before enrollment. Also, they should perform biannual review of their list of medicines and services covered by constantly updating their medicines list according to the pharmaceutical and insurance market.

## INTRODUCTION

There have been several strategies developed globally with a target of improving health financing systems to ensure that people have access to quality health services while being protected from financial burdens which might come from the costs of unforeseen illnesses. One of them being UHC which was introduced in 2012 by the United Nations with the aim of ensuring that all people can obtain the quality health services they need (equity in service use) without fear of financial hardship (financial protection) by expanding the population covered, the package of services, and the extent of financial protection provided (1).

In Tanzania the history of health financing has been a difficult ride. It began with the Arusha Declaration in 1967 whereby free health services for all was introduced under the leadership of the late Julius Kambarage Nyerere, with the aim of removing inequalities in the society and promoting socialism. But in the face of rising costs of health services free health service for all was a failure and the country’s economy could just not sustain the financial burden of health services. Also, in the 1990s health sector reforms that included cost sharing policy was introduced whereby individuals had to contribute and pay for part of the health services offered which was either by risk pooling, or out of pocket money. (2)

Up to December 2019, NHIF’s membership coverage for all stood at 4,856,062 beneficiaries which is equivalent to 9% of the total Tanzanian population. (3)

In the process of increasing health insurance coverage in Tanzania the NHIF rolled out it’s much awaited insurance packages which are designed to make health services more affordable to Tanzanians. In that process three new plans were introduced which were Timiza Afya, Wekeza Afya and Najali Afya whereby every citizen has a chance to enroll in the plans according to his or her needs. The main aim of launching these plans was to target the informal and private sectors which were not covered in the previous NHIF plans.

There are three main packages namely, Wekeza Afya, Timiza Afya and Najali Afya. These packages have been categorized specifically according to one’s economic status, family size, age and the services covered, payments are done once with a one month waiting period.

All the beneficiaries of these package are eligible to acquire services at the level of the dispensary to regional referral hospitals (4).

Therefore, this study evaluated these new packages to close the gap based on the package’s performance and to identify achievements and key challenges. The findings from this evaluation are expected to show the performance of the new packages, by how far have they been able to reach the set target of 50% coverage, level of client satisfactions and health care providers acceptability. This will help to understand the main challenges hindering achieving their goals and best measures which can help to tackle the challenges in order to increase coverage and improve the quality of services offered to its beneficiaries. Also, these findings will be of value as they will act as a baseline for a broader quantitative study which from the findings can help NHIF management team, Ministry of Health Community Development, Gender, Elderly and Children together with other interested stakeholders in decision making and policy formulation towards improving health services in Tanzania.

## METHOD

### Study Context

The study was conducted in health facilities in Kinondoni Municipal and NHIF Kinondoni regional office. Kinondoni Municipal is in northeast of Dar es Salaam Region with a coverage of 531 km^2^ (203 sq. mi) with a population of 1,775,049 people. The municipal has 201 operating health facilities whereby 33 are public health facilities and 168 private health facilities (according PORALG report of 2019). The line of operation of health facilities starts from the Dispensary, Health centre to the Municipal Hospital. The rationale of choosing this study area is because Kinondoni Municipal happens to have the largest number of accredited facilities compared to others and this is according to the Health Facility registry report of 2020(5).

### Study Design

The study used descriptive cross-sectional design using a mixed method approach. Quantitative approach was used to statistically assess the level of utilization of the new packages in terms of number of beneficiaries who have enrolled since the introduction of the packages in September 2019 to March 2021, the time of data collection. Qualitative approach was used to help understanding and interpret beneficiaries and service providers’ perceptions about acceptability and satisfaction towards the benefit of the packages.

### Study Population

The study population included health care providers specifically medical officers because they are the only health care providers allowed to fill in the NHIF consultation claim forms, and NHIF beneficiaries of Timiza, Wekeza and Najali premium packages from the health facilities.

### Sampling

Non-probability sampling was used in this study. NHIF accredited health facilities in Kinondoni Municipal were selected purposively. A total of 18 participants were involved in this study whereby, 9 medical doctors were conveniently selected from the selected accredited health facilities (one public health facility and two private health facilities). Nine (9) beneficiaries of the packages were also conveniently selected from the selected NHIF accredited health facilities. Our sampling was based on saturation principal whereby we continued to recruit participants until no new information was attained from their responses. The facilities were referenced as public or private health facility while maintaining their anonymity.

### Data Collection

In-depth interview guide was used for carrying out the interviews, whereby questions were structured according to the authors experience and literature reviewed. The interviewer defined key concepts of the evaluation so as respondents can be aware of the study. All interviews were conducted in Swahili and each interview lasted between 30 and 45 minutes. During the interview notes were taken by the research assistant who accompanied the PI. Data abstraction tool was used to extract all the members who have enrolled in Najali, Wekeza and Timiza Afya schemes from the beginning of the program September 2019 to the time of data collection in March 2021.

## Data Analysis

Thematic data analysis method was used for analyzing qualitative data which involved reading and re-reading of the data, coding, data reduction, data display and interpretation (6). Data was transcribed and later translated into English language. The English translated data was analyzed through the examination and categorization of respondents’ opinions. The analysis was carried out in four stages. First, the line-by-line coding of field notes and transcripts; Second, themes were searched from the general list of codes created and some codes formed main themes or sub themes, whereas other codes were discarded; third, theme were re red and reviewed at the level of coded data and theme itself to ensure that all the data formed a coherent pattern; fourth themes were then defined and named later final analysis and report writing was done. Data from the abstraction tool was cleaned and imported into Microsoft excel were charts and graphs were constructed to represent the cumulative and the trend of membership enrollment.

### Ethical Considerations

Muhimbili University of Health and Allied Sciences granted ethical clearance for this study (Ref: HD/MUH/T.746/2019). Permission to conduct the study was obtained from National Health insurance Fund, District Medical Officer of Kinondoni and Directors of the hospital and medical officers in charge. The purpose of the study was explained to participants and written consent was sought before the interview. Privacy and confidentiality were highly considered whereby each interviewee was interviewed alone in the room and no names were required. Information provided from the respondents was disclosed for research purposes. Participants were informed that their participation was purely voluntary, and they could withdraw from the study anytime

## Results

The number of members enrolled in the new NHIF packages in Kinondoni Municipal from September 2019 to April 2021 is a total of 17,248 as seen on Figure 2. This number is about 1% of the total population of Kinondoni Municipal which has a total of 1,775,049 people according to the census conducted in 2012. During data collection it was also found that NHIF doesn’t keep records on retainment to track how many members have renewed their memberships. And the data was not disintegrated into the insurance packages as Timiza, Wekeza and Najali.

**Figure 2.**
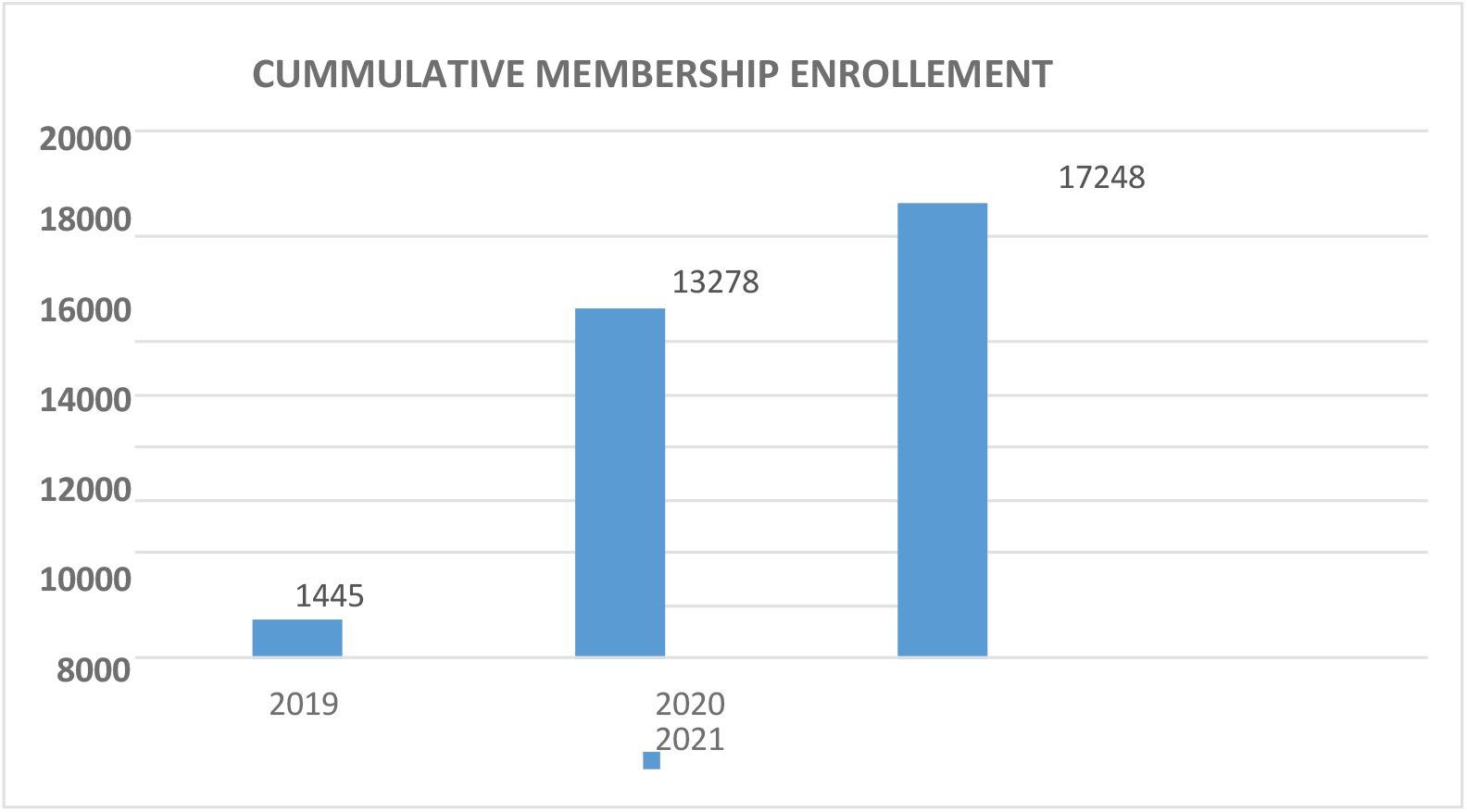
Cumulative enrollment of members from Sept to March 2021 Source: Field data, 2021

### Reduction of out-of-pocket expenditure

Generally, almost all the participants reported to agree that health insurance reduced out of pocket expenditure from medical costs. This is the only sole reason why most of them decide to the join these insurance packages. It was finally an opportunity for them to stop worrying about medical emergencies.

> *“I joined Wekeza Afya Scheme because I am now comfortable that at least when a medical emergency arises, I will be covered with most of services, even if I will be required to top up for some services with out of pocket, the cost won’t be the same as a person not having an insurance at all” (KI-18)*

Another beneficiary reported.

> *“I am now not worried about the costs if I incur our common diseases such as Malaria, UTI and Tonsillitis which previously I used to spend a lot of money on doctor’s consultation and medication” (KI-8)*

Another participant felt that the out-of-pocket expenditure is not reduced at all from her experience.

> *“Since I have joined the scheme, I have always been buying my regular daily medications, as since day one I was told they are not covered, this is a waste of my money. I have to wait for emergency to use my insurance” (KI-04)*.

Despite the findings from the beneficiaries, medical doctors also agreed on the reduction of our pocket expenditure that it has increased number of people who have access to health services. One doctor stated that.

> *“Since the introduction of these packages, the flow of patients has increased and when you try to look at which insurance these patients are in you will find its either Timiza Afya, Najali Afya or Wekeza Afya” (KI-03)*

Another Doctor said.

> *“Honestly these new packages have really helped patients especially during times of emergencies, previously we used to refer these patients to other public facilities due to financial constraints but nowadays the number of referrals from financial constraint has reduced to the minimum” (KI-06)*.

Most of the clients participated expressed a negative attitude towards the services offered by the packages. They feel like it was a trap that NHIF just simply wanted them to enroll in the schemes, but they have not fulfilled their side of the bargain. Most of them expressed that they were not fully oriented on the services covered they mostly came to find out at the time-of-service acquisition in the health facilities. They reported to have lost confidence in the fund, and this might even influence them not to renew their membership for the next year.

### Factors contributing to client dissatisfaction

#### Poor Orientation on the services offered by the packages

All the informants reported that when they went for the first time at NHIF offices for initial registration in the insurance schemes, most of them were just given a paper to read, which had a summary of the price packages, number of investigations, medications and surgical procedures covered. All these services were in numbers and not in the actual service name, therefore most of them selected the packages without knowing if their medications for their conditions are covered or not. One respondent had this to say:

> …*” I choose TIMIZA because it had a higher number of investigations, medications and surgical procedures, being not aware that my medications for my hyperthyroidism are not listed in the package, when you go to NHIF everybody is busy, the staff have no time to sit with you for half an hour and take you through the services before enrollment. I had to pay out of pocket for my monthly medications for the whole year of being a member of TIMIZA Afya; I wasted my money in that insurance”*. (KI-01)

Other informants reported that sometimes the packages are updated or even modified, but the updates are only shared with the health care providers and not them, while they are the primary stakeholders, hence they have all the rights of being updated on any changes concerning the packages. They must find out when they reach the health facilities while sick seeking for services.

> …*” There were some changes early last year concerning the medications covered in the packages, and some hospitals were still refusing to give us some of the medications we do not know if they were doing it for their personal gains or they simply did not have the information on the updates, but we could access the same medication from other hospital which was the same level as the previous one*.*” (KI-02)*

The informants also reported that due to poor orientation and misinformation, during the enrollment process they were only focusing on the prices of the packages and looking forward to choosing a package which is affordable without knowing that they might choose a package which does not cater for their needs. Most of the elderly informants were complaining about their chronic medications which they have been using for a while, and they were never told that they were not covered in their insurance package until they went to the health facilities. One informant had this to say.

> *“*… *My mother has hyperthyroidism, and I am diabetic, the first time I went for enrollment I asked if Vildagliptin and levothyroxine are covered and I was told yes they are covered under Wekeza Afya, but when I went to the hospital, I was told they are not covered in my package” (KI-05)*

#### Cumbersome Referral system

Informants stated that due to the non-transparent and lengthy bureaucratic referral procedures in which they were unaware of which hospitals they can access and which ones they cannot, and on the levels of hospitals categorized by NHIF whereby there are some hospitals allowed to prescribe only a certain medication and some are not. Based on the informant’s experiences many of them complained that they were not told about all these limitations during the time of registration hence they became aware of their existence when visiting the health facilities for treatment. By then they had already paid their insurance premium for the whole year. This uncertainty triggered some of them not wanting to renew their membership because they find it is not useful to them.

> *“*…*the bureaucracy in the referral systems and facility levels of prescription has made me one day visit almost 4 hospitals just looking for Azithromycin every hospital I went were referring me to another one saying that they are not allowed to prescribe it according to the categorized level”. (KI-07)*

Other informants were also concerned about their geographical locations and the hospital they can access according to their packages. Some need to go very far from their homes to seek for medical services just to be referred to a hospital close to where they are living. They noted that most of the times the hospitals which they can access are not operating for 24hours hence bring a lot of inconveniences especially for situations whereby they are supposed to seek for medical attention at night.

> *“*…*We have paid a lot of money for me and my family and there is a hospital closer to where I live but I have to go to a hospital which is about 10km further just to because my package doesn’t allow me to visit the hospital closer to my house without a referral”. (KI-11)*

Doctors had a different perspective about this as all three of them were happy about the referral system, as it had really helped to reduce congestion especially in zonal and national hospitals, as people can get services from health centers and dispensaries before jumping to referral hospitals.

> *“*… *The referral system is very helpful as it really reduces congestion especially for those staff working at zonal and national hospital, cause imagine of every member of these packages could just go to Muhimbili whenever they liked? Things could have been terrible there*.*” (KI-15)*

#### Missing essential medications and investigations in the packages

Concerning availability of essential medications, investigations, and surgical procedures most informants were happy about it, they were able to access all the necessary services they needed, they have never had any challenges concerning the services and most of them are ready to renew their membership. One of them had this to say.

> … *“NHIF has been able to give me my basic need and an opportunity to confidently visit a hospital even if when I don’t have a single cent in my pocket, because with my Wekeza Afya I am sure of getting all the necessary treatment and management without any financial worries, even if I will need to be admitted. I can get admitted in peace without having to worry my family or friends from out-of-pocket expenditure. There are just very few things to be amended concerning the waiting period, I think it should be reduced, but the rest is fine, and I would really like to congratulate the fund and the Government*.*” (KI-12)*

Nine (9) beneficiaries were interviewed 3 from each package. Out of the 9 only 3 said they will renew their schemes; the rest were not satisfied, and they were certain not to renew their membership and the main reason being mostly the availability of medication and investigations.

> *“*… *I am still not sure if am going to renew my Wekeza Afya for next year, to be honest I just feel like I need this insurance for when something bad happens (God forbid) but I am honestly not happy with the services*.*” (KI-09)*

#### Uncertainties

Every medical doctor interviewed had something to say concerning the uncertainties with these packages. It was observed that NHIF seems like they do not educate and take their clients through the packages during enrollment. Most of these clients get to know what is covered or not covered when they visit the health facilities when they are already enrolled and have paid for a full year subscription. This situation brings a lot of commotion and bad reputation of hospitals and practitioners, as clients feel like the doctors/ hospitals have just decided not to give services since they were told all services are available by NHIF.

> *“*… *A patient can come with lower back pain, and as a doctor you know that this patient will benefit from physiotherapy, but since it is not covered you will have to tell the patient that he/she has to pay cash*… *and that is where hell breaks loose, patients will start complaining and tell us that we are thieves, because NHIF has told them everything is covered and they are not to pay cash, so this brings a lot of inconveniences”(KI-10)*

One Doctor also expressed that:

> *“*… *It is uncertain on who is supposed to educate these clients on issues concerning these packages is it NHIF? Or the Doctors working at the hospitals? Because there is not doing his or her job here, and I think is NHIF. They leave the orientation job to us and later we are the ones who suffer from claim rejections” (KI-17)*

From these uncertainties, it was shown that doctors tend to have a bit of a challenge attending clients of Timiza, Wekeza and Najali Afya and hence they prefer attending patients from other NHIF scheme.

But despite that these new packages have shown to increase accessibility to health services to a lot of Tanzanians who could not afford out of pocket expenditures.

> *“*… *These packages have really helped Tanzanians in accessing health services, because we experience cash patients who sometimes come to the hospital severely injured and they cannot even afford a simple x-ray, these kinds of scenarios are also heart breaking to the doctors, but only if this patient had Najali Afya he could have accessed such services even if he had no money” (KI-12)*.

## DISCUSSION

Our findings showed an increase in overall health insurance coverage from the new packages by 1% of the total Kinondoni population which is way below the set target of 50% by June 2020. More than 70% of the total population in 2021 remains uninsured (7). Our findings are comparable with other study reports which showed that more than 60% of citizens in Kenya are also not covered by health insurance (8). The question of why there is poor coverage on health insurance is not unique to Tanzania rather it is an issue in all low- and middle-income countries. Tanzania has a fast-growing informal sector, whose coverage is voluntary. Other studies have also shown that it is hard to achieve UHC with voluntary enrollment mechanism due to unpredictable fluctuating incomes of citizens working in the informal sectors (9). It has been seen that it is hard to track these people for renewing their memberships or even to convince them into joining the schemes. Secondly presumably because according to Tanzania mainland poverty assessment of 2019, almost 50% of Tanzanians live below the poverty line (10), hence they cannot afford the insurance packages introduced. This situation implies that voluntary, contributory approach is unlikely to help in achieving UHC through expanded health insurance coverage. Poverty remains the main factor hindering increasing access to health services.

Among those that were covered by the insurance plans our findings have revealed significant barriers and enablers which might have also contributed on the performance of these health insurance packages.

One of the important enablers was awareness on the importance of having a health insurance. Almost all the informants agreed that they joined the schemes in order to reduce out of pocket expenditure. This finding is like a study in Ghana which showed that the uninsured citizens are more likely to incur catastrophic health expenditures than the insured (11). This has shown that people are aware of the importance of having a health insurance. Despite their level of dissatisfaction with the services, clients believed that being in these schemes was better than not having a health insurance at all. This has in fact somehow increased the number of people who are insured and as a result it has increased the number of people who have access to health services. From the study the 2% of people who have been enrolled into the schemes have somehow reduced their out-of-pocket expenditure which is consistent with findings from other LMIC such as Ghana and Kenya (12).

Among the biggest challenge causing customer dissatisfaction in most of National health insurance schemes is poor coverage of services offered (13). Similar findings were seen in this evaluation. When the packages were first introduced the enrollment, rate was high as people believed that their financial burdens on health costs would be solved, but unfortunately the numbers started declining. This situation might have been caused by clients being dissatisfied with the quality of services hence not referring or recommending others to enroll.

This has partly contributed to the poor enrollment rate, as it is difficult to convince more citizens to enroll in these new schemes while the ones who are already in it are constantly complaining. Beneficiaries are not fully aware of the services offered until when it’s time to go to the hospital and they are denied of certain services that are not covered in their packages.

Not only that but also, the beneficiaries are limited to number of health facilities they can visit. Most of them were not oriented on which hospital they can go, and which they cannot. For those who knew which hospitals to go to complained that most of the selected facilities were far from where they are living. These findings are different from other studies which have shown that insurance companies are working to bring health services closer to the patients by making plans whereby patients can access all health facilities which are closer to client residence. It has also been found that most of public health insurances in other countries have no significance on patient’s choices of health care facility levels (14). Studies have recommended that if insurance schemes are trying to reduce number of people accessing tertiary facilities, hence they should increase the reimbursements of these primary health facilities in order to encourage them to improve their quality of services and hence it will attract patients. Beneficiaries had higher expectations during the time of enrollment as they were made to believe that every health service, they need will be covered but they later got disappointed during the time-of-service acquisition, because they realized that the services are not covered in their insurance package.

If NHIF had time to educate their clients on every service covered and not covered, it would have reduced the number of complaints from the beneficiaries as they will all be comfortable with the packages during the time of enrollment, and they will also know what services to expect and not to expect at the health facilities. In so doing the NHIF would reduce the burden carried by health care providers in explaining service coverage of the packages to the patients during service provision.

Another barrier which has been observed from this evaluation which might have been the root of all evils is poor communication strategy used by NHIF team to their beneficiaries. This finding is congruent to findings of evaluation studies on NHIF by other scholars (16)(15). Poor communication has led to ambiguities on matters such as what is covered and what is not covered in the NHIF packages which has led to patients getting denied of services leading to dissatisfaction. In order to rescue this situation health care providers at the health facilities have been serving as the primary interface between patients and NHIF with respect to their service benefit especially medicines. This has so often served as the main source for customer dissatisfaction towards accredited health facilities on issues which are beyond their control. Poor communication strategy from NHIF towards their beneficiaries has been a setback for them to perform better and this has also been the case in another study done in Tanzania on NHIF relationship with drug outlets (17).

NHIF doesn’t effectively communicate with their clients during the time of enrollment or at any other period of their membership. During the time of enrollment, they simply give them brochures which have different figures of number of services offered and the scheme prices, whereby most of the time clients choose the schemes based on what they can afford instead of choosing based on services covered in the scheme. Secondly, NHIF does not properly communicate benefits changes with its beneficiaries; they only do so with health care providers. This creates confusion and puts health care providers in the position of having to explain changes to the beneficiaries and turn away those who may be unable to afford out of pocket payments. These kinds of situations have put the health care providers in a situation to sometime have poor attitude and acceptability in serving beneficiaries from these packages. This finding is different from other studies such as the evaluation on the Obama Care which have shown that poor acceptability of health care providers towards beneficiaries of National Health Insurance schemes is mostly caused by claim rejections and delay in reimbursements (18).

Every doctor who was interviewed was aware of the existence of the packages, and they also agreed that the packages have increased access to care. However, they expressed their biggest concern about under performance and non-availability of necessary medicines and services which coincides with a study done in districts of Kwa Zulu Natal (19). They agreed that the insurance schemes have been introduced with a good intention of increasing health coverage in the country. But if the services offered are not be disclosed openly to the public during the time of enrollment and only left to the health care providers during consultations, this will scare away citizens from enrolling in the scheme. It is easy for clients to blame the health care providers and health facilities as they are, they are the ones providing the bad news of a service not being covered, hence this partly might bring a negative attitude of providers towards members of these packages. NHIF needs to work on this factor because health care workers acceptability by patients has a direct impact on service utilization and on the quality of care offered in those systems. In many studies findings have shown that job satisfaction of a medical doctor is contributed by quality of services and the extent to which patients feel helped by the medical services provided by the health care provider. These schemes have somehow weakened the autonomy of medical doctors, by increasing their administrative work as now they must memorize which services are covered and which are not covered which was not supposed to be part of their duty. The schemes have also reduced their level of diagnostic accuracy and the level of usage of the medical technology available; they are now obligated to treat these beneficiaries in the lowest possible level of patients’ management.

## CONCLUSION

However, for better performance of the health insurance packages more effort is needed to promote improving their communication process with their beneficiaries and improve relationship with health care providers. They should conduct biannual review of the packages by involving multiple stakeholders including beneficiaries and without forgetting health care providers from all levels of health facilities.

## Data Availability

All data produced in the present study are available upon reasonable request to the authors

## REFERENCES

1. Evans DB. Universal Health Coverage: Concepts and Principles. Heal Syst Finance 2012;(October):1–35.Evans-WHO-UHC-Concepts-and-Principles.pdf

2. Mtei G, Mulligan J, Ally M, Palmer N, Mills A. An Assessment of the Health Financing System in Tanzania: Implication for Equity and Social Health Insurance. Framework.2007;(May).

3. Tanzania, MfukowaTaifaWaBimaYa Afya. https://www.nhif.or.tz. 2019. p. 1 https://www.nhif.or.tz/pages/profile#gsc.tab=0

4. FomuyaUsajiliWanachamawaVifurushi NHIF 1F.pdf. https://www.nhif.or.tz/uploads/publications/en1587454644-FomuyaUsajiliWanachamawaVifurushiNHIF1F.pdf

5. Ntundu RA. Process Evaluation of Nhif Service Provision in Accredited Health Facilities: A Case of Temeke Municipal Health Facilities.1999.

6. LaxmaiahManchikanti M, Standiford Helm II M, Ramsin M. Benyamin M, Joshua A. Hirsch M. A Critical Analysis of Obamacare: Affordable Care or Insurance for Many and Coverage for Few? Pain Physician [Internet],2017

7. Linje GO and R. Customer satisfaction with national health insurance fund services: a case study of selected public and private hospitals in Moshi municipality, tanzania.2005.

8. Ministry of health, community development, gender elderly and children. Tanzania health facility registry (hfr) public portal [Internet].

9. Vaismoradi M, Jones J, Turunen H, Snelgrove S. Theme development in qualitative content analysis and thematic analysis. J Nurs Educ Pract.2016;6(5).

10. Kazungu JS, Barasa EW. Examining levels, distribution and correlates of health insurance coverage in Kenya. Trop Med Int Heal.2017;22(9):1175–85.

11. Okoroh J, Essoun S, Seddoh A, Harris H, Weissman JS, Dsane-Selby L, et al. Evaluating the impact of the national health insurance scheme of Ghana on out-of-pocket expenditures: A systematic review. BMC Health Serv Res.2018;18(1).

12. The World Bank. Mainland Poverty Assessment. Tanzania Main Poverty Assessment.2019.

13. Wang H, Zhang D, Hau Z, Yan F, Hou Z. Association between social health insurance and choice of hospitals among internal migrants in China: A national cross-sectional study. BMJ Open.2018;8(2).

14. Latiff-Khamissa S, Naidoo P. Knowledge, awareness and readiness of private sector doctors practicing in the Ethekweni and Ugu districts of KwaZulu-Natal province for the implementation of the National Health Insurance. South African Fam Pract.2016;58(1):18–23

